# “Now you see me”: detecting asymptomatic infectious individuals in the population

**DOI:** 10.1101/2023.05.08.23289685

**Authors:** Alena Iseli, Akos Dobay, Lutz Philipp Breitling

## Abstract

Quarantine is an effective countermeasure to stop or slow the spread of an emerging infectious disease when no other preventive measures are available to protect the population. However, when the disease results in a proportion of asymptomatic infections, the spread dynamics are affected, and quarantine efficiency is impaired.

Here, we introduce an extended susceptible-infected-recovered (SIR) model to study the effects of asymptomatic individuals at the onset of an emerging infectious disease when no vaccination is yet available and/or when a vaccine is available but only a subset of the population can be vaccinated due to limited supply or the unwillingness of susceptible individuals to receive an injection. These aspects have been indirectly incorporated into the model using a time-dependent vaccination rate.

With this model, we confirm that, in the case of a missing vaccine, quarantine is effective in stopping the spread of an infectious disease, but its efficiency can be substantially reduced in the presence of individuals developing asymptomatic infection. Moreover, we show that vaccination is effective only if available early during the epidemic and if the vaccination rate is sufficiently high. By applying this model to Zurich and all of Switzerland in case of the COVID-19 pandemic, we found that the following two strategies have similar outcomes: either placing infectious individuals into quarantine when no vaccine is available or dropping quarantine measures but administering a vaccine at a daily rate of 1%, starting no later than 105 days after the onset of the epidemic. Beyond this time period, a vaccination campaign will have no effect in stopping the spread of the disease if 25% of the susceptible population is asymptomatic. We also found that the option of deploying a vaccination campaign was more effective for all of Switzerland than for only the city of Zurich.

## 1. Introduction

In December 2019, the city of Wuhan in Hubei Province, China, experienced an outbreak of pneumonia of unknown etiology. While the initial hope was to contain the disease, later defined as COronaVIrus Disease 2019 (COVID-19), the rapid spread of the virus gave rise to a new pandemic [1]. The spreading of severe acute respiratory syndrome coronavirus 2 (SARS-CoV-2), the causative agent of COVID-19, was preceded by two other epidemics in the 21^st^ century caused by viruses of the same family: SARS-CoV in 2002 and 2003 and Middle East respiratory syndrome coronavirus (MERS-CoV) in 2012 [2].

An effective way of preventing contact with sick people and therefore avoiding a potential infection is to keep them isolated. Quarantine or isolation has a long history and can be traced back to biblical times. During the Renaissance, when trade between Europe and Asia was booming, the cities of Venice and Genoa were two powerful maritime republics. Venice used quarantine (from *quarantena*, meaning forty, and before it, the *trentino*, or thirty, first imposed in 1347 in the Republic of Ragusa, Dalmatia, in modern Dubrovnik (Croatia)) [3] for any ship entering the harbor before the crew and passengers were allowed to disembark and the crew to unload the cargo. The principle underlying quarantine was that, if a person was infected with a disease, a period of forty days was sufficient to display the symptoms, preventing an infectious individual from potentially starting an outbreak. Since then, quarantine measures have been used worldwide in various circumstances and situations. In some cases, quarantine can take the form of self-isolation as an extremely tight measure to control the spread of an infectious disease, and sometimes it provides the only possibility of eradicating a deadly infectious disease. Such extreme measures can be attested in Europe. An example is the plague outbreak between 1582 and 1583 at the port of Alghero in Sardinia [4]. Under the guidance of the physician Quinto Tiberio Angelerio, the city introduced prophylactic measures, including self-isolation and lockdowns, to contain the epidemic [4]. Many of the measures present during the 1582-83 outbreak at the port of Alghero remind us of the social distancing measures adopted by most of the countries around the world to fight the COVID-19 pandemic [5].

Quarantine is an efficient measure to prevent the outbreak of an infectious disease, but it can badly fail if susceptible individuals develop an asymptomatic infection or if the detection measures are inadequate [6]. To understand how asymptomatic individuals can affect the spread of an emerging or re-emerging infectious disease, we propose studying a modified version of the basic susceptible-infected-recovered (SIR) model. The SIR model was introduced in the first half of the 20^th^ century by the Scottish biochemist William O. Kermack (1898-1970) and the Scottish military physician and epidemiologist Anderson G. McKendrick (1876-1943), and it was published in three separate papers [7, 8, 9]. The mathematical model uses the same principle of a chemical reaction by subdividing the population into groups, each having a well-defined state. In the case of an infectious disease, each individual in the population will undergo a transformation that will change them from being in a susceptible (S) state into an infected (I) state and finally into a recovered or removed (R) state. The number of individuals in each group at a given time point and their subsequent transitions from one into the other state are described by a set of nonlinear, first-order ordinary differential equations (ODEs). In this description, the variation in the number of infected individuals is directly proportional to the product of the number of susceptible and infected individuals. The proportionality factor is called the *force of infection*. Hence, encounters between a healthy (susceptible) and an infectious individual may lead to a new infection. The model description also includes the *recovery rate*, defined as the inverse of the time period during which an individual is infectious. The rate at which infected individuals are infecting other susceptible individuals in the population is provided by the *basic reproduction number R*_0_ (pronounced “R naught”, where the subscript zero stands for the time *t* = 0). If *β* represents the infection rate or the daily number of newly infected individuals in the population, *γ* the recovery rate per capita, and *S*_0_ the actual number of susceptible individuals in the population at the start of an epidemic, the value of *R*_0_ is given by

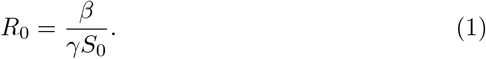

The value of *R*_0_ corresponds to the basic reproduction number at the start of an epidemic. This number can be recomputed at any subsequent time point to account for the actual dynamics of the epidemic. In the literature we also find the *force of infection λ* as the rate of infection per capita [10]. Using 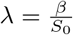 instead of *β* we can write

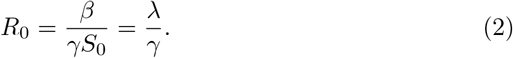

Extended SIR models for SARS-CoV-2 epidemics were largely investigated in 2020, giving rise to a large body of literature [11, 12, 13, 14, 15]. In particular, Dobrovolny developed an extended SIR model that specifically includes a distinction between symptomatic and asymptomatic individuals and their effects on the current SARS-CoV-2 epidemic in the US [16]. In our extended SIR model we divided the susceptible population into symptomatic and asymptomatic individuals. In this context, the words “symptomatic” and “asymptomatic” describe a natural tendency of the individual to display or not to display symptoms, and not the individual’s health state. This model allows us to study the relative proportions of asymptomatic individuals in the population and their effect on the spread of the disease. In addition, the model includes: (1) quarantine countermeasures; and (2) vaccination.

As an example, we propose to look at the COVID-19 pandemic in a city with the size of Zurich and, for comparison on a larger scale, in a country such as Switzerland.

## 2. Materials and Methods

### 2.1. Overview of the model

How a healthy individual will react to a newly released pathogen spreading across a population is unknown. To incorporate this information into a basic SIR model, we divided the population into two subgroups: susceptible individuals who would develop a symptomatic infection upon contracting the disease (*S*_1_); and those who would experience an asymptomatic infection (*S*_2_). Hence, the overall population of susceptible individuals is the sum of the two proportions *S*_1_ + *S*_2_. Using this assumption, we can write

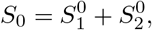

and with this notation, our definition of *R*_0_ becomes

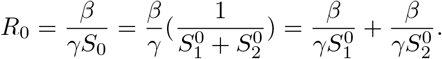

This division allows us to analyze the dynamics of an epidemic by varying *S*_1_ and *S*_2_ in a dependent manner and by examining the effect of their relative frequency on the spread of the disease. As a consequence of this partition, the time progressions of *S*_1_ and *S*_2_ individuals throughout the epidemic remain separate. The model also enables the study of different recovery rates, assuming that symptomatic infected individuals seeking medication earlier than asymptomatic infected individuals might also recover faster. For convenience and whenever possible, we write (*S*_1_ + *S*_2_) as *S*_1,2_. The other variables in the model are the infected symptomatic and asymptomatic individuals (*I*_1_ and *I*_2_, or *I*_1,2_), the quarantined infected individuals (*Q*_1_ and *Q*_2_, or *Q*_1,2_), the vaccinated susceptible individuals (*V*_1_ and *V*_2_, or *V*_1,2_), and the recovered individuals (*R*_1_ and *R*_2_, or *R*_1,2_). Figure 1 shows the flow diagram of the modified version of the basic SIR model. The set of ODEs (Equation 2) describes the state variables and their respective transition rates.

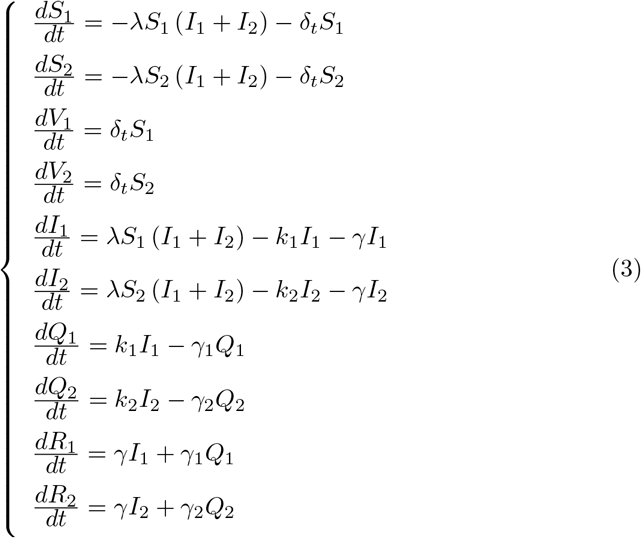

**Figure 1.**
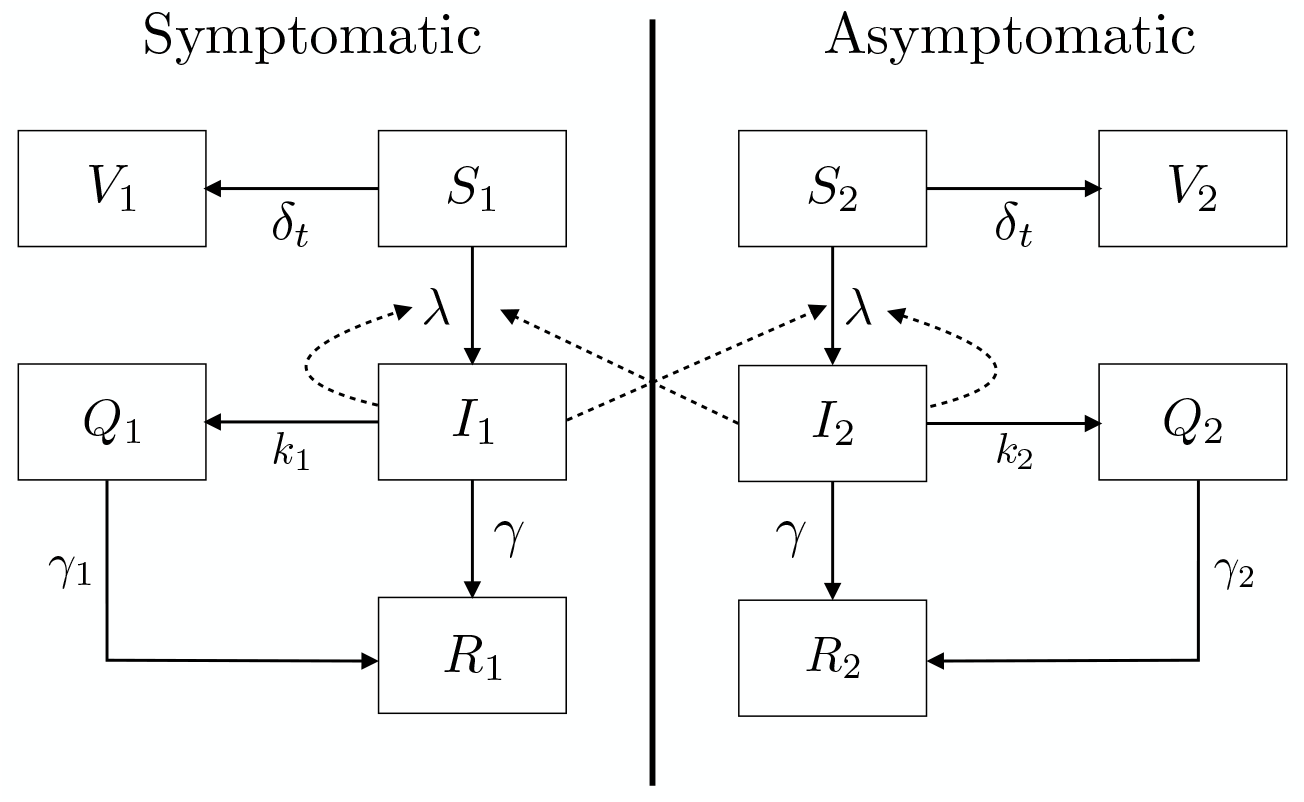
Flow diagram of the extended SIR contact model showing the evolution of an infectious disease when the population is constituted by subgroups of symptomatic (*S*_1_) and asymptomatic (*S*_2_) susceptible individuals. The model controls for (self)-isolation (*k*_1_ and *k*_2_) and time-dependent vaccination rates (*δ*_*t*_). The other variables are infected individuals (*I*_1_ and *I*_2_), quarantined infected individuals (*Q*_1_ and *Q*_2_), vaccinated susceptible individuals (*V*_1_ and *V*_2_), and recovered individuals (*R*_1_ and *R*_2_).

In addition to *λ*, we have *δ*_*t*_, the daily vaccination rate: it describes the proportion of vaccinated susceptible individuals removed from the pool of susceptible individuals. For simplicity, we assume that only susceptible individuals can be vaccinated. The values *k*_1_ and *k*_2_ define the daily quarantine rates, i.e., the proportion of infected individuals removed from the pool of infectious individuals. In contrast to the historical quarantine regulations, which were usually applied to all individuals as preventive measures, we focus on measures that concern only infected individuals (i.e., individuals who tested positive for a pathogen, with or without displaying symptoms). The values *γ, γ*_1_, and *γ*_2_ describe the various daily recovery rates. In contrast to the quarantine rates *k*_1_ and *k*_2_, *δ*_*t*_ is a time-dependent parameter. It is equal to zero if the considered time point *t* is strictly less than a predefined threshold *θ*, and it takes a defined value *δ*_*θ*_, strictly greater than zero, when *t* is greater than or equal to *θ*:

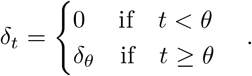

This property was chosen because a vaccine is usually not readily available when a new disease emerges. Furthermore, we can assume that, for an epidemic, the population size remains more or less constant, and

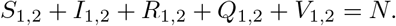

In the special case when

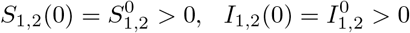

and

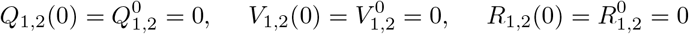

One can derive an expression for the daily relative removal rate *ρ*, the rate at which the infected population subgroup varies in size, and the basic reproduction number *R*_0_ [17]. The expression can be obtained by solving *dI*_1_*/dt* on one side and *dI*_2_*/dt* on the other side. Hence, for *t* = 0, we have:

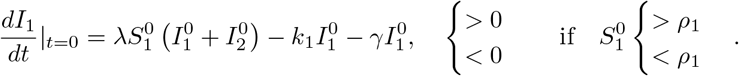

The relative removal rate for symptomatic individuals becomes

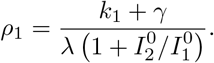

A similar expression can be derived for *dI*_2_*/dt*

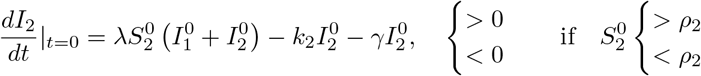

leading to the relative removal rate for asymptomatic individuals:

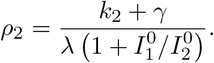

The basic reproduction number, *R*_0_, can be derived from the relative removal rate since an epidemic starts when

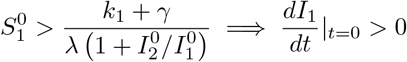

or when

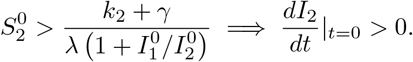

In the first case, the contribution of the symptomatic susceptible people to the basic reproduction number is

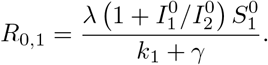

In the second case, due to the symmetry of our model, we have

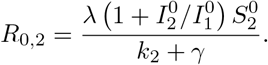

Hence, the overall contribution of both symptomatic and asymptomatic individuals results in a basic reproduction number of

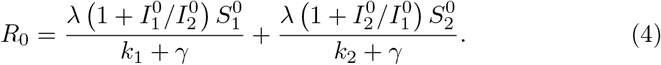

### 2.2 Model assumptions

In this model, we assumed the same probability (included in *λ*) of being infected by either *I*_1_ or *I*_2_. If this assumption is not the case, a slight modification of the equations is necessary. Additionally, symptomatic individuals (*S*_1_) are assumed to (self)-isolate faster than asymptomatic individuals (*S*_2_), resulting in *k*_1_ being strictly larger than *k*_2_ (*k*_1_ *> k*_2_). This outcome can be achieved by either staying at home or in a healthcare facility. Consequently, infectious symptomatic individuals (*I*_1_) remain in the population for a shorter period of time and thus infect fewer people. As a comparison, we also included the case in which *k*_1_ is equal to *k*_2_ (*k*_1_ = *k*_2_). Regarding the recovery rate, the model offers some flexibility, but it is fair to assume that *γ*_1_ and *γ*_2_ are strictly greater than *γ* (*γ*_1_, *γ*_2_ *> γ*). In other words, staying at home or in a healthcare facility should lead to a shorter recovery period. Similarly, we can assume that, in the case of an effective treatment, *γ*_1_ is strictly greater than *γ*_2_, and both are strictly greater than *γ* (*γ*_1_ *> γ*_2_ *> γ*): assuming that a faster (self)-isolation/(self)-medication can lead to faster recovery. Finally, one can set *γ*_1_ equal to *γ*_2_ and *γ* (*γ*_1_ = *γ*_2_ = *γ*). The parameters *β, λ, γ*, and *δ*_*t*_ are assumed to be the same for symptomatic and asymptomatic patients, but this assumption can easily be modified.

An additional assumption for this model is that, at the beginning of the epidemic, all individuals are equally susceptible to infection. This assumption is reasonable since the population is largely immunologically naïve against an emerging disease. In our investigation, the incubation time following the infection, before becoming infectious, was neglected. Finally, we assume that the probability of recovery or the recovery rate (*γ*) is constant and that the infection confers lifelong immunity. For vaccination, a single-dose vaccine that provides immediate and long-lasting immunity is assumed. Although some of the aforementioned assumptions are major, we intentionally chose them to keep the model simple and to specifically study the effects of quarantine and isolation. However, changes to these assumptions can be made to increase the model complexity.

### 2.3. Implementation

The model was implemented in Python. The code was adapted from “Learning Scientific Programming with Python” [18] and is available on GitHub.

## 3. Results and Discussion

To study our model, we simulated various epidemic scenarios. As starting parameters, we used the population size of Zurich, the largest city in Switzerland, and we extended the simulations countrywide to all of Switzerland. To assess the evolution of the disease in Zurich (Figures 2A, 2C, 3A, and 3C) and in the country (Figures 2B, 2D, 3B, and 3D), we plotted the phase trajectories of the epidemic using the total susceptible (*S*_1,2_) or total susceptible and vaccinated individuals (*S*_1,2_ + *V*_1,2_) versus total infected (*I*_1,2_) individuals. For all trajectories in Figures 2 and 3, one symptomatic infected individual (patient zero) was placed into the whole population of susceptible individuals 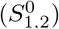. The value of *R*_0_ was set to 2.5, corresponding to an early estimate by the WHO in the context of the COVID-19 outbreak [19]. The choice of *γ* = 1*/*11 is based on the median time required by asymptomatic individuals and patients with a noncritical symptomatic infection to clear SARS-CoV-2, as determined by Kumar and colleagues [20]. The value of *λ* was different depending on 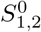. The trajectories were determined over a time period of 365 days.

**Figure 2.**
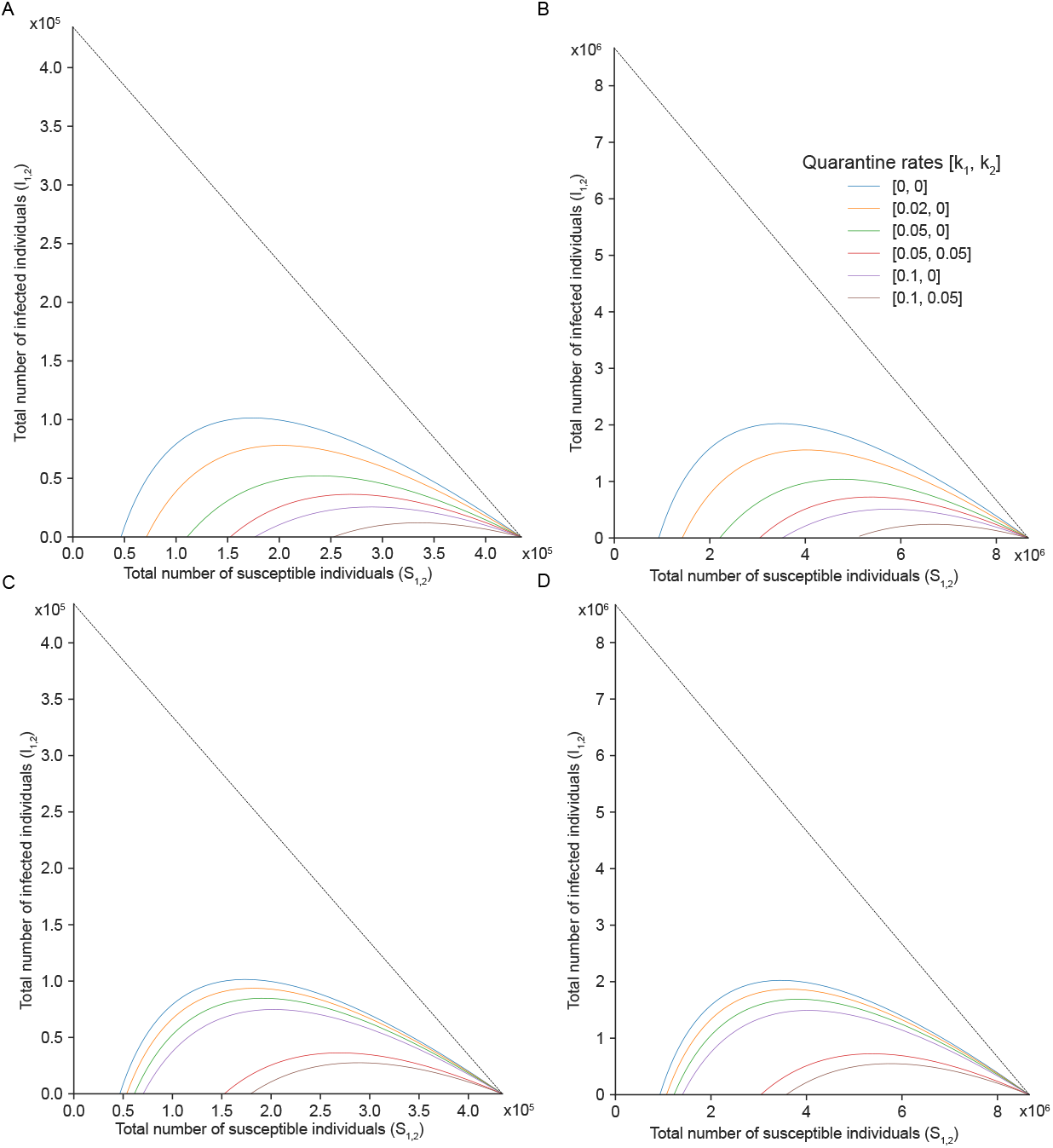
A population with 25% (A and B) and 75% (C and D) asymptomatic individuals (*S*_2_). Phase trajectories in the total susceptible individuals (*S*_1,2_) versus total infected (*I*_1,2_) individuals plane with no vaccination (*δ*_*t*_ = 0). All the trajectories are determined by *γ* = 1*/*11, and *δ*_*t*_ = 0. The different trajectories correspond to different values of the quarantine rates (*k*_1_, *k*_2_). No quarantine corresponds to the case in which [*k*_1_, *k*_2_] = [0, 0], while a fast rate corresponds to [*k*_1_, *k*_2_] = [0.1, 0.05]. Given the initial number of susceptible individuals, the force of infection, and the recovery rate, the basic reproduction number (without quarantine) is *R*_0_ = 2.5. The plots are determined by the following conditions: (A) the city of Zurich *N* = 434, 536, 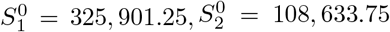, *λ* = 5.23 *·* 10^−7^; (B) Switzerland *N* = 8, 667, 100, 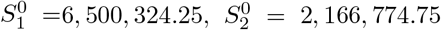, *λ* = 2.62 *·* 10^−8^; (C) the city of Zurich *N* = 434, 536, 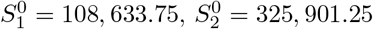, *λ* = 5.23 *·* 10^−7^; (D) Switzerland *N* = 8, 667, 100, 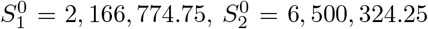, *λ* = 2.62 *·* 10^−8^. The remaining values are 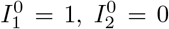, and 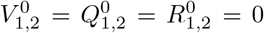. The black dotted line defines the total number or individuals *N*.

**Figure 3.**
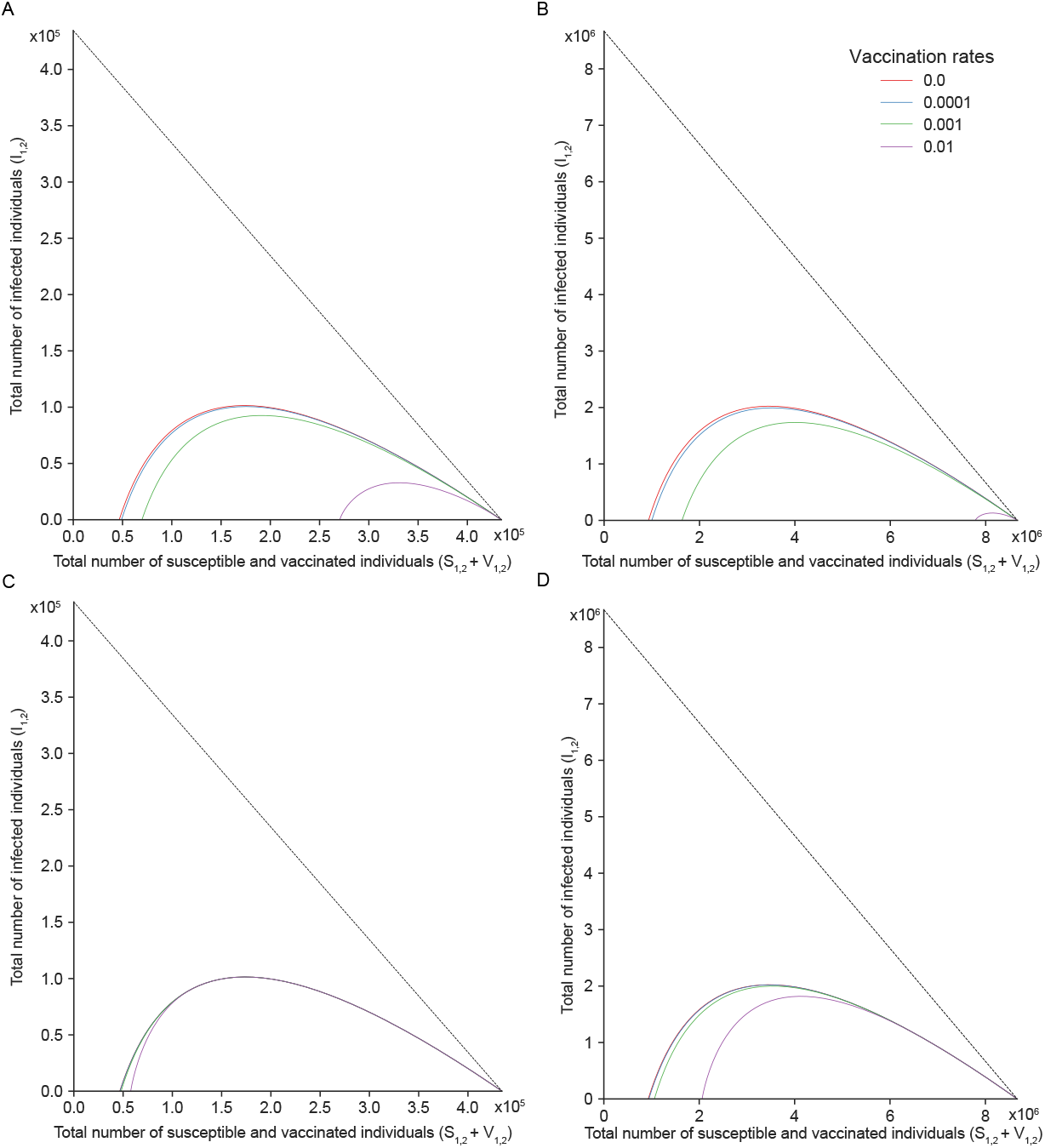
Effects of vaccination. Phase trajectories in the total susceptible and vaccinated individuals (*S*_1,2_ + *V*_1,2_) versus total infected (*I*_1,2_) individuals plane with vaccination. All the trajectories are determined by *γ* = 1*/*11, using 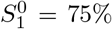 and 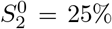. The different trajectories correspond to different vaccination rates after 50 days (A and B) and 105 days (C and D). The quarantine rate was set to zero, corresponding to a basic reproduction number of *R*_0_ = 2.5. The plots are determined by the following conditions: A) and C) the city of Zurich *N* = 434, 536, 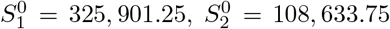, *λ* = 5.23 *·* 10^−7^; B) and D) Switzerland *N* = 8, 667, 100, 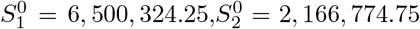, *λ* = 2.62 *·* 10^−8^. The remaining values are 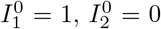, and 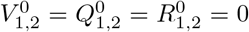. The black dotted line defines the total number or individuals *N*.

In the first scenario, the parameters for quarantine and vaccination were set to zero, implicating that *Q*_1,2_ and *V*_1,2_ remained empty during the epidemic (Figure 2 with [*k*_1_, *k*_2_] = [0, 0], blue line, and Figure 3 with *δ*_*t*_ = 0 for any *t >* 0, red line). This scenario shows the size of the epidemic in the absence of any interventions. Figure 2 shows the size of the epidemic in Zurich (2A) and Switzerland (2B) when the population is predominantly composed of symptomatic individuals 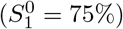. The various trajectories in the susceptible-infected phase space highlight the effects of quarantine from low (*k*_1_ = 0.02) to high (*k*_1_ = 0.1). A quarantine rate of *k*_1_ = 0.05 or 5% applied to symptomatic individuals while leaving asymptomatic individuals without isolation is sufficient to reduce the size of the epidemic by half (see also Supplementary Table 1). In comparison, when the population is dominated by asymptomatic individuals 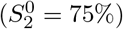, the peak of the epidemic is only decreased by approximately 16% for [*k*_1_, *k*_2_] = [0.05, 0] compared to 50% in the previous case (Figure 2C and 2D; see also Supplementary Table 1). Quarantine rates of [*k*_1_, *k*_2_] = [0.1, 0.05] are sufficient to prevent or reduce the size of an epidemic both in Zurich and in Switzerland. These results emphasize the impact of asymptomatic individuals in a population in which quarantine is the only measure available to counteract the spread of an emerging disease. When asymptomatic individuals are highly prevalent in a population, test campaigns are important to detect false negatives, i.e., infected individuals who do not display any symptoms.

In the second scenario, in the absence of isolation, we investigated the effects of vaccination by introducing different vaccination rates between 0 (*δ*_*t*_ = 0) and 1% (*δ*_*t*_ = 0.01), starting 50 days after the beginning of the epidemic (Figure 3A and 3B), and 105 days after the beginning of the epidemic (Figure 3C and 3D). In this scenario, we assumed that the population had a natural tendency toward symptomatic individuals. To be consistent with the first scenario presented in Figure 2, we decided to split the population according to the same proportion with 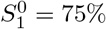 and 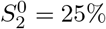. Similar to the quarantine scenario, vaccinations cause the number of individuals who never became infected to increase (in this scenario *S*_1,2_ + *V*_1,2_) at the end of the epidemic when compared to the scenario with no state intervention. However, the time point of vaccine administration is critical: when susceptible individuals can be inoculated starting 50 days after the beginning of the epidemic, the proportion of susceptible and vaccinated individuals (*S*_1,2_ + *V*_1,2_) at the end of the epidemic can be as high as approximately six times the value corresponding to the case when no vaccination is available at the city level and eight times the value at the country level (see also Supplementary Table 2). This scenario would be the case with a 1% vaccination rate (*δ*_*t*_ = 0.01). The longer it takes to deploy a vaccination campaign, the less efficient it is. In our scenario, we looked at multiple time points and we concluded that, beyond 105 days, the efficacy of a vaccine to prevent the spread of the disease is almost irrelevant in the case of a city, while on a larger scale, such as for Switzerland, approximately 20% of the population does not become infected with *δ*_*t*_ = 0.01.

Interestingly, in our example, the impact of quarantine does not differ between cities and countries, while for vaccination, a difference can be observed. This outcome can be explained by the temporal shift of the infection peak: at the country level, the maximal number of infected individuals is reached later than at the city level. This discrepancy can be explained by the population density: Zurich has a higher population density than Switzerland, with 926.8 [21] persons per km^2^, while Switzerland has only 219 [22] persons per km^2^. This difference is reflected in the force of infection *λ* (see the figure captions for more details). Since the vaccination rate *δ*_*t*_ is a time-dependent parameter, different phase trajectories are observed. According to these plots, vaccination can prevent the disease from spreading if administered early and at high rates. Examples from other infectious diseases show, for instance, how the timing of vaccination campaigns in reaction to measles outbreaks constitutes a critical parameter [23, 24, 25, 26]. However, there are several limiting factors that arise when vaccination campaigns are deployed. First, a vaccination campaign must comply with preliminary steps from the early development phase to clinical trials. Second, it depends on its approval by regulatory agencies. Additionally, it must be distributed on a large scale, and healthcare professionals are needed to inoculate the population. Furthermore, the rate depends on the willingness of the population to be vaccinated, which is partly influenced by the information made available to the public. Finally, one must consider that not all types of vaccines provide full protection; i.e., vaccinated individuals can still become infected. Depending on the efficiency of the vaccine, several administrations might be necessary, with both organizational and economical consequences.

Quarantine, conversely, relies on how effectively local authorities can implement such measures. This action depends largely on resource availability, how local authorities can inform the population, and the willingness of the population to comply with measures. Compared to a vaccine, which might require additional research and developments, quarantine provides a faster solution. Hence, quarantine might be the most efficient way to control the spread of an emerging disease, together with an effective detection rate, when vaccination is not yet available.

Finally, it is worth stressing that it is extremely difficult to obtain exact numbers about how many people have been infected by SARS-CoV-2 during the pandemic. However, the choice of a basic reproduction number of *R*_0_ = 2.5 is in agreement with values available for SARS-CoV-2.

## 4. Conclusion

We developed an extended SIR model to study the size of an epidemic when the population is constituted of symptomatic and asymptomatic individuals: susceptible individuals who have a natural tendency to show no symptoms when they are infected with a particular disease. With this model, we could show the effect of asymptomatic susceptible individuals and how they could affect quarantine measures. Any asymmetries in the ratio of symptomatic/asymptomatic individuals have impacts. In the absence of vaccination campaigns, asymptomatic susceptible individuals remain an issue that can only be controlled indirectly using periodic testing methods in the population, engaging the population to perform self-testing upon known contact with infected individuals, or the wearing of preventive measures, such as personal masks. While this strategy might be considered infringing on individual freedom and could constitute an unnecessary burden in everyday life if it has to be repeated over a long period of time, it offers the advantage of preventing more restrictive measures with a higher economic impact, such as lockdowns. Therefore, a proper detection strategy for asymptomatic infections is instrumental for fighting the spread of a disease.

## Data Availability

All data produced in the present work are contained in the manuscript

## References

[1] C. Wang, P. Horby, F. Hayden, G. Gao, A novel coronavirus outbreak of global health concern, Lancet 395 (10223) (2020) 470–473. doi:10.1016/S0140-6736(20)30185-9. URL https://doi.org/10.1016/S0140-6736(20)30185-9

[2] Y. Yang, F. Peng, R. Wang, K. Guan, T. Jiang, G. Xu, J. Sun, C. Chang, The deadly coronaviruses: The 2003 sars pandemic and the 2020 novel coronavirus epidemic in china, Journal of autoimmunity 309 (2020) 102434. doi:10.1016/j.jaut.2020.102434. URL https://doi.org/10.1016/j.jaut.2020.102434

[3] P. Mackowiak, P. Sehdev, The origin of quarantine, Clinical Infectious Diseases 35 (9) (2002) 1071–1072. doi:10.1086/344062. URL https://doi.org/10.1086/344062

[4] R. Bianucci, O. Benedictow, G. Fornaciari, V. Giuffra, Quinto tiberio angelerio and new measures for controlling plague in 16th-century alghero, sardinia, Emerging Infectious Diseases 19 (9) (2013) 1478–1483. doi:10.3201/eid1909.120311. URL https://doi.org/10.3201/eid1909.120311

[5] The 432-year-old manual on social distancing, https://www.bbc.com/future/article/ 20210107-the-432-year-old-manual-on-social-distancing, accessed: 2021-09-15.

[6] A. Dobay, G. Gall, D. Rankin, H. Bagheri, Renaissance model of an epidemic with quarantine, Journal of Theoretical Biology 317 (2013) 348–358. doi:10.1016/j.jtbi.2012.10.002. URL https://doi.org/10.1016/j.jtbi.2012.10.002

[7] W. Kermack, A. McKendrick, Contribution to the mathematical theory of epidemics, Proceedings of the Royal Society of London. Series A, Containing Papers of a Mathematical and Physical Character 115 (772) (1927) 700–721. doi:10.1098/rspa.1927.0118. URL https://doi.org/10.1098/rspa.1927.0118

[8] W. Kermack, A. McKendrick, Contributions to the mathematical theory of epidemics, part ii, Proceedings of the Royal Society of London. Series A, Containing Papers of a Mathematical and Physical Character 138 (1932) 55–83. doi:10.1098/rspa.1932.0171. URL https://doi.org/10.1098/rspa.1932.0171

[9] W. Kermack, A. McKendrick, Contributions to the mathematical theory of epidemics, part iii, Proceedings of the Royal Society of London. Series A, Containing Papers of a Mathematical and Physical Character 141 (1933) 94–112. doi:110.1098/rspa.1933.0106. URL https://doi.org/10.1098/rspa.1933.0106

[10] M. J. Keeling, P. Rohani, Modeling Infectious Diseases in Humans and Animals, 1st Edition, Princeton University Press, Princeton, N.J., 2007. URL http://amazon.com/o/ASIN/0691116172/

[11] J. Wangping, H. Ke, S. Yang, C. Wenzhe, W. Shengshu, Y. Shanshan, W. Jianwei, K. Fuyin, T. Penggang, L. Jing, L. Miao, H. Yao, Extended sir prediction of the epidemics trend of covid-19 in italy and compared with hunan, china, Frontiers in Medicine 7 (2020) 169. doi:10.3389/fmed.2020.00169. URL https://www.frontiersin.org/article/10.3389/fmed.2020.00169

[12] K. B. Law, K. M. Peariasamy, B. S. Gill, S. Singh, B. M. Sundram, K. Rajendran, S. C. Dass, Y. L. Lee, P. P. Goh, H. Ibrahim, N. H. Abdullah, Tracking the early depleting transmission dynamics of covid-19 with a time-varying sir model, Scientific Reports 10 (1) (2020) 21721. doi:10.1038/s41598-020-78739-8. URL https://doi.org/10.1038/s41598-020-78739-8

[13] V. Grimm, F. Mengel, M. Schmidt, Extensions of the seir model for the analysis of tailored social distancing and tracing approaches to cope with covid-19, Scientific Reports 11 (1) (2021) 4214. doi:10.1038/s41598-021-83540-2. URL https://doi.org/10.1038/s41598-021-83540-2

[14] V. Deo, G. Grover, A new extension of state-space sir model to account for underreporting – an application to the covid-19 transmission in california and florida, Results in Physics 24 (2021) 104182. doi:https://doi.org/10.1016/j.rinp.2021.104182. URL https://www.sciencedirect.com/science/article/pii/S2211379721003302

[15] K. B. Law, K. M. Peariasamy, B. S. Gill, S. Singh, B. M. Sundram, K. Rajendran, S. C. Dass, Y. L. Lee, P. P. Goh, H. Ibrahim, N. H. Abdullah, Tracking the early depleting transmission dynamics of covid-19 with a time-varying sir model, Scientific Reports 10 (1) (2020) 21721. doi:10.1038/s41598-020-78739-8. URL https://doi.org/10.1038/s41598-020-78739-8

[16] H. M. Dobrovolny, Modeling the role of asymptomatics in infection spread with application to sars-cov-2, PLoS ONE 15 (8) (2020) e0236976.

[17] J. D. Murray, Mathematical Biology. I. An Introduction, Vol. 17 of Interdisciplinary Applied Mathematics, Springer, Berlin and Heidelberg, 1993.

[18] C. Hill, Learning scientific programming with python, Cambridge University Press doi:10.1017/CBO9781139871754. URL https://doi.org/10.1017/CBO9781139871754

[19] Coronavirus disease 2019 (covid-19) situation report- 46, https://www.who.int/docs/default-source/coronaviruse/situation-reports/20200306-sitrep-46-covid-19.pdf?sfvrsn=96b04adf.

[20] N. Kumar, A. AbdulRahman, S. AlAli, S. Otoom, S. L. Atkin, M. AlQahtani, Time till viral clearance of severe acute respiratory syndrome coron-avirus 2 is similar for asymptomatic and non-critically symptomatic individuals, Frontiers in Medicine (Lausanne) 8 (616927).

[21] Population in zurich, https://knoema.com/atlas/Switzerland/Zurich/Population-density, accessed: 2021-12-24.

[22] Population in switzerland, https://www.worldometers.info/world-population/switzerland-population, accessed: 2021-12-24.

[23] R. F. Grais, X. D. Radiguès, C. Dubray, F. Fermon, P. J. Guerin, Exploring the time to intervene with a reactive mass vaccination campaign in measles epidemics, Epidemiology and Infection 134 (4) (2006) 845–849. doi:10.1017/S0950268805005716.

[24] C. Dubray, A. Gervelmeyer, A. Djibo, I. Jeanne, F. Fermon, M.-H. Soulier, R. F. Grais, P. J. Guerin, Late vaccination reinforcement during a measles epidemic in niamey, niger (2003-2004)., Vaccine 24 (18) (2006) 3984–3989. doi:10.1016/j.vaccine.2006.01.049.

[25] R. F. Grais, A. J. K. Conlan, M. J. Ferrari, A. Djibo, A. Le Menach, O. N. Bjørnstad, B. T. Grenfell, Time is of the essence: exploring a measles outbreak response vaccination in niamey, niger., J R Soc Interface 5 (18) (2008) 67–74. doi:10.1098/rsif.2007.1038.

[26] M. J. Ferrari, R. F. Grais, N. Bharti, A. J. K. Conlan, O. N. Bjørnstad, L. J. Wolfson, P. J. Guerin, A. Djibo, B. T. Grenfell, The dynamics of measles in sub-saharan africa, Nature 451 (7179) (2008) 679–684. doi: 10.1038/nature06509. URL https://doi.org/10.1038/nature06509

