## Supplemental Table 1 and Table 2 for "“Now you see me”: detecting asymptomatic infectious individuals in the population"

Table .1: Values corresponding to the plots in Figure 2

| $S_0$ | $S_1$ (%) | $k_1$ | $k_2$ | $I_{\max}$ | $S_\infty$ |
|---|---|---|---|---|---|
| 434,536 | 75 | 0.0 | 0.0 | 101,456.83 | 46,649.25 |
| 434,536 | 75 | 0.02 | 0.0 | 78,088.11 | 71,324.27 |
| 434,536 | 75 | 0.05 | 0.0 | 52,074.32 | 110,734.28 |
| 434,536 | 75 | 0.05 | 0.05 | 36,335.84 | 152,589.18 |
| 434,536 | 75 | 0.1 | 0.0 | 25,571.15 | 176,339.05 |
| 434,536 | 75 | 0.1 | 0.05 | 12,086.26 | 252,638.76 |
| 434,536 | 25 | 0.0 | 0.0 | 101,456.83 | 46,649.25 |
| 434,536 | 25 | 0.02 | 0.0 | 93,645.19 | 53,592.44 |
| 434,536 | 25 | 0.05 | 0.0 | 84,747.32 | 61,449.86 |
| 434,536 | 25 | 0.05 | 0.05 | 36,335.84 | 152,589.18 |
| 434,536 | 25 | 0.1 | 0.0 | 74,800.68 | 70,340.36 |
| 434,536 | 25 | 0.1 | 0.05 | 27,575.03 | 179,237.28 |
| 8,667,100 | 75 | 0.0 | 0.0 | 2,023,452.23 | 930,458.33 |
| 8,667,100 | 75 | 0.02 | 0.0 | 1,557,062.24 | 1,422,622.28 |
| 8,667,100 | 75 | 0.05 | 0.0 | 1,039,033.53 | 2,208,694.33 |
| 8,667,100 | 75 | 0.05 | 0.05 | 724,502.53 | 3,043,544.48 |
| 8,667,100 | 75 | 0.1 | 0.0 | 510,017.02 | 3,518,863.71 |
| 8,667,100 | 75 | 0.1 | 0.05 | 241,027.32 | 5,088,619.01 |
| 8,667,100 | 25 | 0.0 | 0.0 | 2,023,477.38 | 930,458.34 |
| 8,667,100 | 25 | 0.02 | 0.0 | 1,867,662.38 | 1,068,945.86 |
| 8,667,100 | 25 | 0.05 | 0.0 | 1,689,720.90 | 1,225,668.69 |
| 8,667,100 | 25 | 0.05 | 0.05 | 724,521.27 | 3,043,544.45 |
| 8,667,100 | 25 | 0.1 | 0.0 | 1,491,957.78 | 1,402,997.96 |
| 8,667,100 | 25 | 0.1 | 0.05 | 549,970.32 | 3,575,271.43 |

Table .2: Values corresponding to the plots in Figure 3

| $S_0$ | $S_1$ (%) | $k_1$ | $k_2$ | $\delta_t$ | $t$ (day) | $I_{max}$ | $(S+V)_\infty$ |
|---|---|---|---|---|---|---|---|
| 434,536 | 75 | 0.0 | 0.0 | 0 | 50 | 101,456.83 | 46,649.25 |
| 434,536 | 75 | 0.0 | 0 | 0.0001 | 50 | 100,558.06 | 48,951.65 |
| 434,536 | 75 | 0.0 | 0.0 | 0.001 | 50 | 92,581.02 | 69,758.96 |
| 434,536 | 75 | 0.0 | 0.0 | 0.01 | 50 | 32,855.40 | 270,537.18 |
| 434,536 | 75 | 0.0 | 0.0 | 0 | 105 | 101,456.83 | 46,649.25 |
| 434,536 | 75 | 0.0 | 0.0 | 0.0001 | 105 | 101,456.83 | 46,787.62 |
| 434,536 | 75 | 0.0 | 0 | 0.001 | 105 | 101,456.83 | 48,000.88 |
| 434,536 | 75 | 0.0 | 0.0 | 0.01 | 105 | 101,456.83 | 57,782.69 |
| 8,667,100 | 75 | 0.0 | 0.0 | 0 | 50 | 2,023,477.38 | 930,458.33 |
| 8,667,100 | 75 | 0.0 | 0.0 | 0.0001 | 50 | 1,994,294.08 | 999,659.84 |
| 8,667,100 | 75 | 0.0 | 0.0 | 0.001 | 50 | 1,735,582.62 | 1,633,281.30 |
| 8,667,100 | 75 | 0.0 | 0.0 | 0.01 | 50 | 130,789.92 | 7,783,945.59 |
| 8,667,100 | 75 | 0.0 | 0.0 | 0 | 105 | 2,023,477.38 | 930,458.33 |
| 8,667,100 | 75 | 0.0 | 0.0 | 0.0001 | 105 | 2,020,985.01 | 943,663.79 |
| 8,667,100 | 75 | 0.0 | 0.0 | 0.001 | 105 | 1,998,765.88 | 1,060,613.15 |
| 8,667,100 | 75 | 0.0 | 0.0 | 0.01 | 105 | 1,818,371.10 | 2,060,998.95 |
